# All-cause mortality during SARS-CoV-2 Pandemic in India: Nationally-representative estimates independent of official death registry

**DOI:** 10.1101/2021.07.20.21260577

**Authors:** Sabareesh Ramachandran, Anup Malani

## Abstract

We estimate excess deaths in India during the COVID pandemic using monthly deaths in the sample of a privately-conducted, nationally-representative, large, panel data set. The data set includes roughly 174,000 households (1.2 million members) and spans January 2015 - June 2021. We estimate COVID is associated with 3.36 million (95% CI: 2.08-4.63 million) excess deaths, a 17.3% increase in the all-cause death rate, until April 2021. Excess deaths spike during the peaks of the 2 infection waves in India. The second wave is associated with significantly more excess deaths than the first. The age-pattern of deaths is skewed towards the elderly relative to baseline.

## Introduction

India has reported 30.7 million SARS-CoV-2 infections and 404,211 deaths through July 7, 2021^1^. However, India’s death registration system does not capture all deaths, from COVID or otherwise^2, 3^. We examine deaths reported independently of that system from a large, nationally-representative, panel data set of roughly 174,000 households (1.2 million persons) from January 1, 2015 - June 30, 2021 to estimate excess deaths during the pandemic.

## Methods

Data are drawn from the Centre for Monitoring Indian Economy’s Consumer Pyramids Household Survey (CPHS). All panel households are surveyed every 4 months (a “round”); a representative 1/4 are surveyed each month. Households report member deaths since the last survey. We assign deaths reported in month t to, on average, have occurred in month t-(k/2), where k is months since the last completed survey. Because timing of deaths is more reliable among households that respond in consecutive rounds, we focus on such households.

We estimate an individual-level linear regression of whether a member died in a month on a constant and indicators either for each month, for the pandemic period (March 2020 - April 2021), or for each wave (wave 1, March-December 2020; wave 2, January-April 2021). The constant estimates the baseline death rate. The coefficient on time-period estimates the excess death rate during that time period. To estimate demographic-specific rates, we include indicators for these groups and the interaction of those indicators with the pandemic indicator. Non-response weights are used to address non-response in each survey round.

We primarily employ data from January 2019-May 2021, so our baseline period is January 2019- February 2020. However, there was a jump in death rates in the CPHS in 2019, as in other sources^4^. Therefore, we also report estimates when the baseline period is January 2015- February 2020.

We test for differences in coefficients with 2 sided t-tests. Statistical analysis was conducted with Stata v16.1.

## Results

Response rates averaged 85% until 2020. Response rates fell to 65%, 45% (due to India’s lockdown), 72%, and 74% in the 4 rounds starting January 2020.

Baseline mortality is 0.79% (95% CI: 0.77 - 0.81%) in 2015-2019 and 1.07% (95% CI: 1.03 - 1.11%) in 2019. Mortality rose starting May 2020, with peaks in September 2020 and April 2021 (Figure 1). Using 2019 as a baseline, average excess death rate was 0.185% (95% CI: 0.115 - 0.255%), 0.119% (95% CI: 0.039 - 0.199%), and 0.336% (95% CI: 0.229 - 0.444%) during the pandemic, wave 1 and wave 2, respectively. Wave 2 had significantly higher excess mortality (p < 0.001).

**Figure 1.**
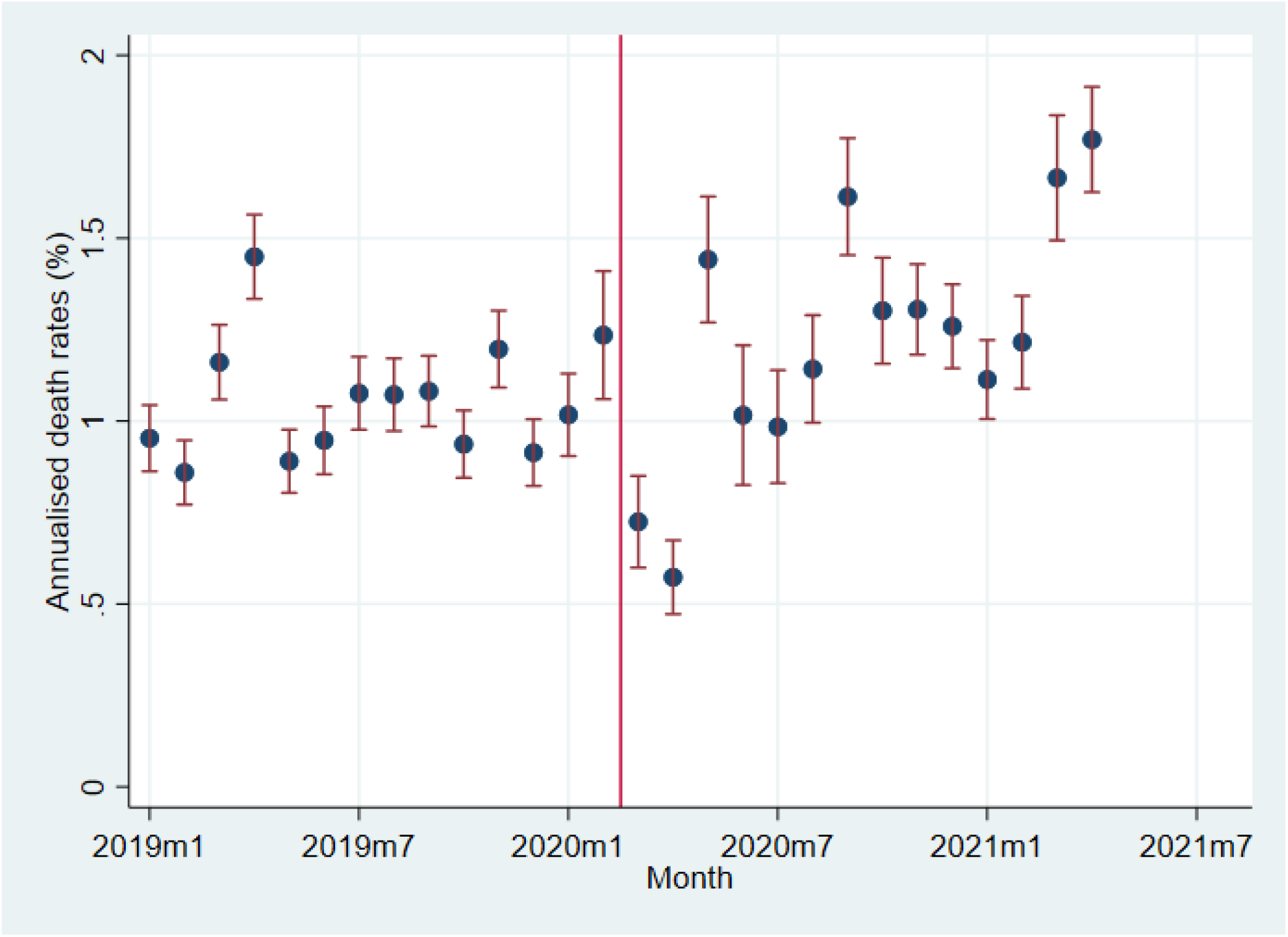
Estimated excess death rate by month. Notes. Plot of coefficients on month indicators from regression of whether a person dies on month indicators. The coefficients have been multiplied by 300 to show annualized death rates in percent. Sample includes data from January 2015 - May 2021, though only data from 2019 onwards are plotted. Only individuals that respond to consecutive surveys are included. Individual death reported in month t attributed to month t-2.

Excess death rate was significantly higher relative to 2019 baseline for ages 60-69 and 70+ (p < 0.001) (Figure 2).

**Figure 2.**
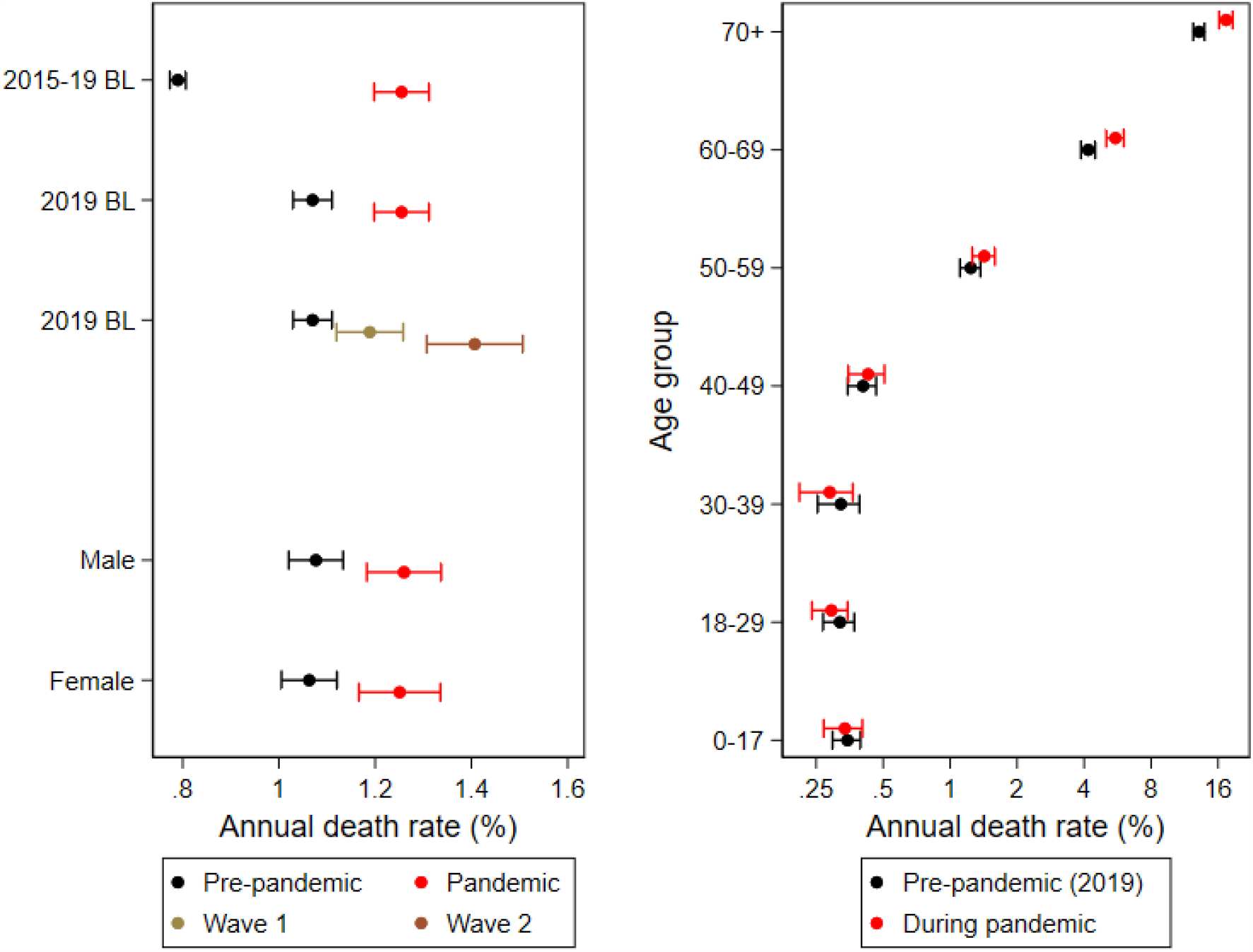
Plot of estimated all-cause death rates during pandemic, by wave, sex and age. Notes. Each row presents a plot of coefficients and 95% confidence intervals on the constant, pandemic or wave indicators and on the interaction of pandemic and demographic group.

## Discussion

COVID increased the all-cause death rate by 17.3%, compared to 22.9% in the US ^5^. The estimated excess deaths is 3.36 million (95% CI: 2.08-4.63 million), implying 8.4 times more people died than official COVID deaths through June 2021. The timing of observed deaths matches the peaks of India’s 2 waves. The age pattern of deaths matches that of COVID.

Our findings have several limitations. First, response rates were low, especially during India’s lockdown. However, much of the non-response was because CPHS randomly sampled only 50% of households during the lockdown. We employ non-response weights to adjust non-response. Moreover, non-responding households in one round may report deaths from that round in subsequent rounds; we estimate excess deaths using that data in the Supplement.

Second, CPHS may not be as representative as official excess-death numbers ^6^. We estimate a higher baseline death rate in 2019 than Global Burden of Disease ^4^. However, official all-cause mortality is only available for 10 states (out of 28) and the two sources give comparable answers.

## Online Supplement

### Data

The CPHS is a proprietary data set. Licenses can be obtained by any member of the public at https://consumerpyramidsdx.cmie.com/.

Table S1 presents 3 types of response rates for each round. One is the response rate among all households that round. The second is the percent of households that respond in that round and the subsequent round. The third is the percent of households that respond in that round or in some subsequent round. Note that estimates for the second rate is missing for round 1 2021, as that is the last date of our data; the third rate converges to the first rate by round 1 2021 because we do not have data on responses in future rounds after the last round.

### Methods

To determine if non-response is random, we estimate two sets of regressions. Observations are at the household x month level. The first set is intended to test whether CPHS genuinely selected households at random to visit during lockdown. Initially, focus on months prior to 2020, before the pandemic. The dependent variable is whether CPHS contacted a household in a round. The independent variables are chosen by LASSO from the hundreds of variables on each household and each member of each household in the CPHS (https://consumerpyramidsdx.cmie.com/). After selecting regressors from pre-2020 data in this manner, we switch the sample to months in 2020 and 2021. We estimate a regression where the dependent variable is whether CPHS contacted a household in a round, and the independent variables are the regressors chosen in the previous regression. We record the R2 from regressions on each of these samples. The final regressions repeat the last two regressions, but switch the samples.

The second set of regressions is intended to test whether households contacted by CPHS respond at random. Initially, focus on rounds prior to 2020, before the pandemic. The dependent variable is whether a household contacted by CPHS responded to the survey. The independent variables are chosen by LASSO from the hundreds of variables on each household and each member of each household in the CPHS (https://consumerpyramidsdx.cmie.com/). After selecting regressors from pre-2020 data in this manner, we switch sample to rounds in 2020 and 2021. We estimate a regression where the dependent variable is whether a household CPHS contacted in a round responded to the survey, and the independent variables are the regressors chosen in the previous regression. We record the R2 from regressions on each of these samples. The final regressions repeat the last two regressions, but switch the samples.

Finally, to address the problem of low response rate to consecutive surveys, we estimate a version of the regression in the main text including households that do not respond in consecutive rounds because, as Table S1 shows, the response rate (rate 3) is higher among this group than in the sample including only households that respond in consecutive months (rate 2).

### Results

The R^2^ in the household-contacted regression is very low (< 0.04) whether the regressions are picked by LASSO for the pre-pandemic period or for the pandemic period. The R2 in the household-responded regression is also very low (< 0.04) whether the regressions are picked by LASSO for the pre-pandemic period or for the pandemic period.

Across consecutive- and non-consecutive responding households, baseline mortality is 0.86% (95% CI: 0.85 - 0.88%) in 2015-2019 and 1.12% (95% CI: 1.08 - 1.16%) in 2019. Mortality rose starting May 2020, with peaks in October 2020 and May 2021 (Figure S1). Using 2019 as a baseline, average excess death rate was 0.247% (95% CI: 0.18 - 0.31%), 0.233% (95% CI: 0.16 - 0.30%), and 0.295% (95% CI: 0.18 - 0.41%) during the pandemic, wave 1 and wave 2, respectively. The difference between Waves 1 and 2 is not statistically significant.

### Discussion

CPHS appeared to select the subsample of households to contact during lockdown and the remainder of the pandemic at random conditional on observable variables in CPHS. Likewise, the subsample of households contacted by CPHS during lockdown and the remainder of the pandemic appear to respond at random conditional on those observable variables.

That said, it is possible selection of households to contact or household response decisions could be non-random but correlated with unobservable variables. However, the CPHS has a very large number of measurements on demographics, income, time use, labor market status and consumption; therefore, there may not be very much unobservable and concerning variation available to explain contact and response rates. Moreover, even this large number of observable variables explains a very small percent of variation in those rates. It is unlikely there is substantial non-random non-response unless concerning unobservable variables explain more variation in rates than observable variables.

Using data even from households that do not respond in consecutive rounds, the estimated excess deaths during the pandemic is 5.21 million (95% CI: 4.06-6.36 million), implying 13 times more people died than official COVID deaths through June 2021. The timing of observed deaths again matches the peaks of India’s 2 waves. The age pattern of deaths matches that of COVID.

## Data Availability

The Centre for Monitoring Indian Economy's Consumer Pyramids Household Survey data are a proprietary data set. Licenses can be obtained by any member of the public at https://consumerpyramidsdx.cmie.com/.

## Tables and Figures

**Table S1.**
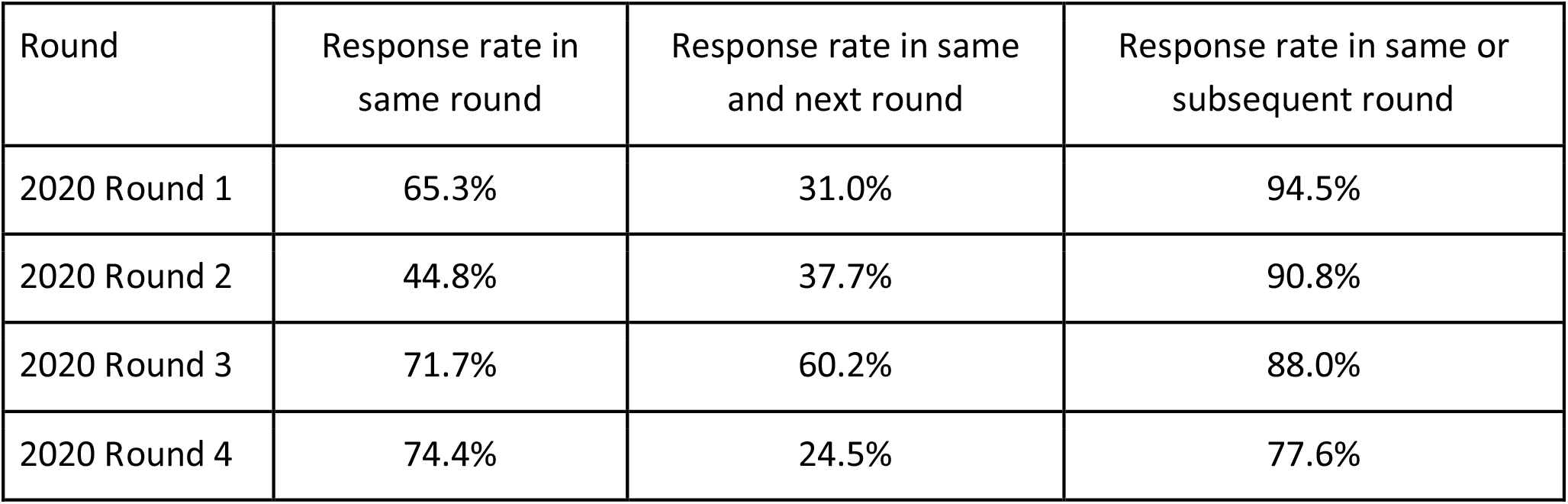
Response rate by round, defined as response current or subsequent rounds.

| Round | Response rate in same round | Response rate in same and next round | Response rate in same or subsequent round |
| --- | --- | --- | --- |
| 2020 Round 1 | 65.3% | 31.0% | 94.5% |
| 2020 Round 2 | 44.8% | 37.7% | 90.8% |
| 2020 Round 3 | 71.7% | 60.2% | 88.0% |
| 2020 Round 4 | 74.4% | 24.5% | 77.6% |

**Figure S1.**
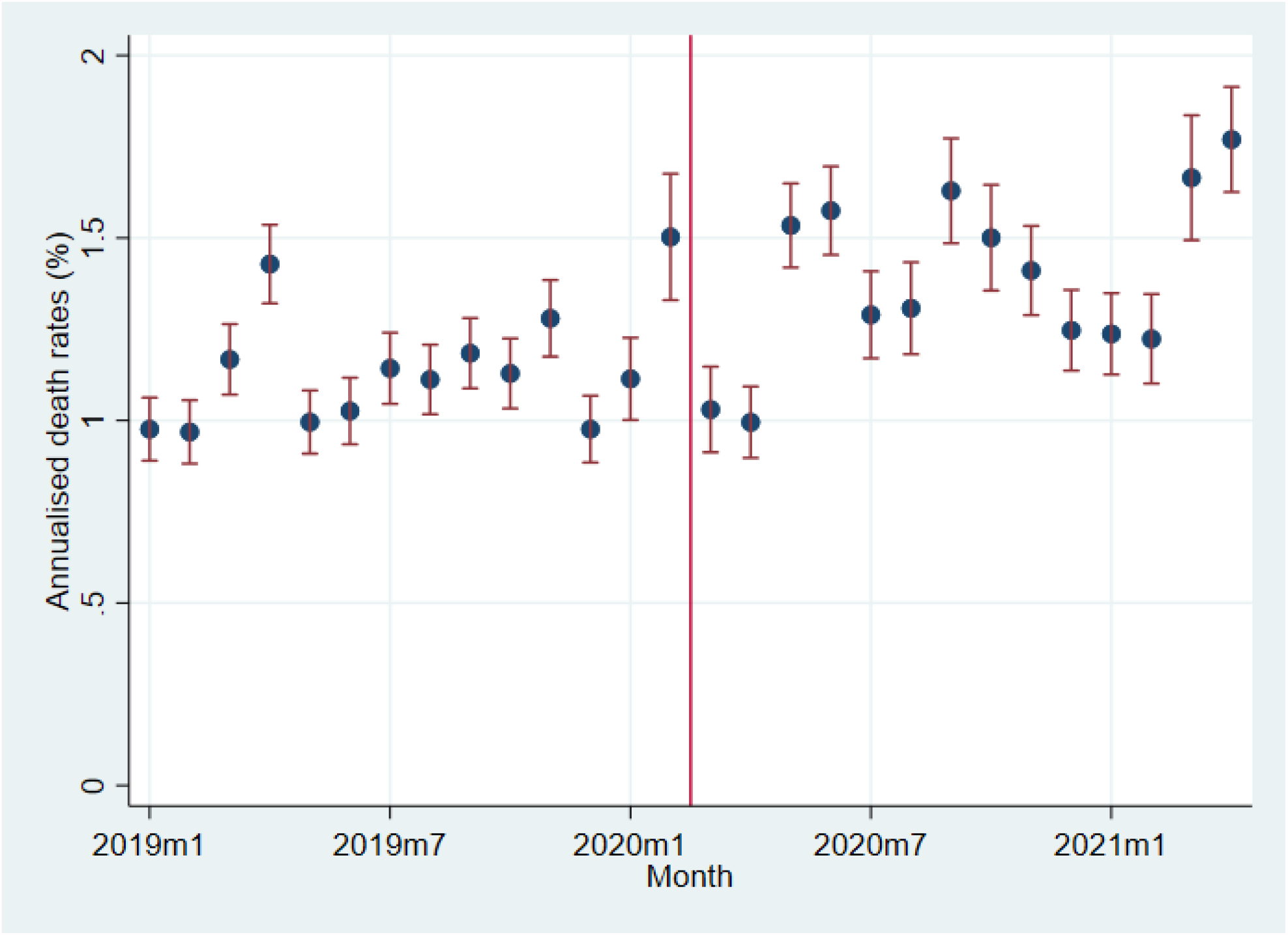
Estimated excess death rate by month, using consecutive and non-consecutive responders. Notes. Plot of coefficients on month indicators from regression of whether a person dies on month indicators. The coefficients have been multiplied by 300 to show annualized death rates in percent. Sample includes data from January 2015 - May 2021, though only data from 2019 onwards are plotted. Individuals that respond to consecutive and non-consecutive surveys are included. Individual death reported in month t attributed to month t-(k/2), where t-k is the last month in which the individual responded to a survey.

## Acknowledgments

Access to the CPHS database employed in this study was funded by the Becker Friedman Institute at the University of Chicago. The funder had no role in design and conduct of the study; collection, management, analysis, and interpretation of the data; preparation, review, or approval of the manuscript; and decision to submit the manuscript for publication. Malani (University of Chicago) and Ramachandran (University of California San Diego) had full access to all the data in the study and take responsibility for the integrity of the data and the accuracy of the data analysis.

We are grateful to Chinmay Tumbe for comments on this research.

